# Detection of Common Deletion Mutations (− α^3.7^ and − α^4.2^ kb) in *HBA* gene and Genotype-Phenotype Correlation

**DOI:** 10.1101/2024.09.03.24312976

**Authors:** Satarupta Basu, Soma Gupta, Rajib De, Shuvra Neel Baul, Aditi Sen, Shreyashi Dasgupta, Arindam Biswas

## Abstract

**Background and Objectives:** Microcytic hypochromic anemia is the most common feature of alpha-thalassemia and depends on the number of alpha genes deleted. Therefore, in this study, we aim to determine the most common deletion mutations among microcytic anemia cases of West Bengal and correlate them with different biochemical parameters and endophenotypes.

**Methods:** Two hundred and sixty patients with microcytic anemia were recruited. The GAP-PCR technique was adopted to identify the 3.7 kb and 4.2 kb deletion mutation in the *HBA* gene.

**Results:** Forty patients were found to have either 3.7 kb or 4.2 kb deletion mutations which represents ~15.38% of the microcytic anemia patient population. A statistically significant lowering of MCH values (P = 0.02) and elevated levels of Total Bilirubin (P = 0.0001), direct bilirubin (P = 0.0004), unconjugated bilirubin (P = 0.0001), SGPT (P = 0.005), SGOT (P = 0.005), ALP (P = 0.008), RBC count (P = 0.01) and Hemoglobin level (P = 0.02) were observed among the alpha-thalassemia mutation carriers than non-mutant. The study failed to show any correlation between mutation status disease severity and gender bias.

**Conclusion:** The two common deletion mutations (−3.7 kb or -4.2 kb) in the *HBA* gene are most commonly found in microcytic anemia cases of West Bengal and can be correlated with several biochemical parameters.

## INTRODUCTION

Thalassemia is one of the most common inherited autosomal recessive blood diseases in the world, characterized by absent or decreased production of normal hemoglobin, resulting in microcytic anemia of varying degrees. There are two major types of thalassemia, α- and β-, which are caused by mutations in the α- and β-globin genes respectively. Beta-thalassemia is extremely prevalent in India with a carrier frequency ranging from 7-10% in the general population [Sharma, 1963; Misra, 1991; Dash, 1985] and over 500 different mutations in the *HBB* gene have been reported in different parts of the world. Mutations in *HBA1* and *HBA2* genes on chromosome 16 will result in alpha-thalassemia. Individuals with a single α-globin gene mutation are silent carriers, with two mutations known as alpha-thalassemia traits that may display mild anemia, and three mutations that are affected require health monitoring. Alpha-thalassemia may occur due to deletional or non-deletional mutations in the α-globin gene, and deletion mutations are more common. The most commonly identified single deletion mutations in Southeast Asia are – α^3.7^ and – α^4.2^ kb deletion. Apart from these, other deletion variants are also found among the different world populations which include – α^20.5^, --^SEA^, --^MED^, --^THAI,^ and --^FIL^ [Karakas, 2015]. Approximately, 5-20% of the world’s population carries either one or more alpha-thalassemia mutations. In India, the overall prevalence of alpha-thalassemia was 12.9% and highest among the Punjabi population originating from the northern region of India [Nadkarni, 2008]. Sen et al, reported that the *alpha-globin* gene deletion alleles were found in 18% of subjects from West Bengal, 3.9% from Arunachal Pradesh, and 3.84% from the tribal population of Assam and co-existence of *alpha-* and *beta-globin* gene mutations were observed up to 18% of the tribal population [Sen, 2005]. It was also documented that there is a significant difference in mean corpuscular volume (MCV; P<0.001) and mean corpuscular hemoglobin (MCH; P<0.001) among the Indonesia alpha-thalassemia deletion mutation carriers [Husan, 2021]. Therefore, in this study, we aimed to detect the two common (– α^3.7^ and – α^4.2^ kb) alpha-thalassemia deletion mutations using the GAP-PCR technique and correlate them with the haematological, biochemical, and clinical parameters among the microcytic anemia population of West Bengal.

## MATERIALS AND METHODS

### Recruitment of study subjects

Patients attending the Haematology OPD clinic at Nil Ratan Sircar Medical College and Hospital, Kolkata, West Bengal, India were screened for microcytic hypochromic anemia based on Complete Blood Count (CBC). Two hundred and sixty patients (age range 2-82 years) with MCV <80 fl, MCH < 27pg were recruited in this study to estimate iron profile, liver function test, genetic analysis, etc. The demographic details are mentioned in Table 1. All subjects provided written informed consent as per the guidelines of the Indian Council of Medical Research (ICMR). The Institutional Ethics Committee on research using human samples cleared the project after proper review.

**Table 1:** Demographic details of study subjects according to *alpha-thalassemia* gene mutations.

| **Subjects/Genotypes** | **Male** | **Female** | **Age Range (years)** |
| --- | --- | --- | --- |
| Study subjects (n = 260) | 123 (47%) | 137 (53%) | 2 – 82 |
| $\alpha\alpha/\alpha\alpha$ (n = 220) | 104 (47%) | 116 (53%) | 2 – 82 |
| $\alpha\alpha/\alpha$-3.7 (n = 30) | 14 (47%) | 16 (53%) | 3 - 60 |
| α-3.7/α-3.7 (n = 7) | 4 (57%) | 3 (43%) | 3 - 44 |
| α-3.7/α-4.2 (n = 1) | 1 (100%) | 0 | 18 |
| $\alpha\alpha/\alpha$-4.2 (n = 1) | 0 | 1 (100%) | 26 |
| α-4.2/α-4.2 (n = 1) | 0 | 1 (100%) | 60 |
| Allele frequency (-3.7 kb deletion mutation) | 23 (4.67%) | 22 (4.01%) | NA |
| Allele frequency (-4.2 kb deletion mutation) | 1 (0.203%) | 3 (0.547%) | NA |

#### Collection of blood samples and genomic DNA preparation

2 mL of peripheral blood samples were collected in EDTA vials from patients. Genomic DNA was extracted from all study subjects using QIAamp DNA Blood Mini Kit (Qiagen GmbH, Hilden, Germany) according to the manufacturer’s instructions and dissolved in TE (10 mM Tris− HCl, 0.1 mM EDTA, pH 8.0) and stored at 4^0^C.

### Haematological Analysis

3 mL of peripheral blood samples collected in plain vials and separated serum was estimated for LFT and Iron profile. Among LFT, the Concentration of total and conjugated bilirubin (Unconjugated bilirubin was calculated), Total protein, and albumin were estimated. Enzyme activity, namely AST (Aspartate transaminase or SGOT, ALT (Alanine transaminase or SGPT) and alkaline phosphatase were determined. So far Iron profile is concerned, Total iron concentration and unbound iron binding capacity (UIBC) were measured. Total iron binding capacity was calculated using a standard formula.

All parameters were estimated using an autoanalyser (XL 600, Transasia Biomedicals Ltd).

#### GAP-Polymerase chain reaction (PCR) and Agarose Gel Electrophoresis

To detect the two common deletion mutations (−α3.7 and -α4.2 kb deletion) in the *HBA* gene, the GAP-PCR approach was taken. In this method, a single-tube multiplex PCR assay is capable of detecting any combination of -α3.7 and -α4.2 kb deletion mutations in the *HBA* gene. Primers were designed to amplify the junction fragments of the α-thalassemia determinants that could be easily identified by size (Chong et al., 2000).

#### Statistical analysis

Biochemical parameters and comparison of clinical parameters were analyzed by unpaired T-test and Fischer exact P-value.

## RESULTS

### Genetic analysis

Two hundred and sixty patients with a mean age of 36.77±19.74 (age range 2-82 years) were recruited in this study. A total of 40 patients were found to have either 3.7 kb or 4.2 kb deletion mutations (Table 1), which represents ~15.38% of the patient population. Out of forty individuals, thirty individuals were heterozygous for α3.7 deletion (−α3.7/αα), seven samples were homozygous for α3.7 deletion (−α3.7/-α3.7), one sample was heterozygous for both -α3.7 and -α4.2 deletion (−α3.7/-α4.2), one sample was heterozygous for 4.2 kb deletion (αα /-α4.2), one sample was homozygous for 4.2 kb deletion (−α4.2/-α4.2) and the remaining 220 were normal (αα/αα) for these deletion mutations screened. We did not find any differences in mutation frequency between male and female cases (15.45% in males and 15.32% in females). In our study cohort, ~4.71% (49/1040) of deleted alleles (either 3.7 kb deletion or 4.2 kb deletion) were found of which 91.8% of 3.7kb deletion and 8.2% of 4.2 kb deletion mutation. The 3.7kb and 4.2kb deletion mutations were present among 14.62% (38/260) and 1.15% (3/260) of the study subjects respectively.

### Comparison of biochemical parameters

Next, we compared the biochemical parameters like Bilirubin, SGOT, SGPT, ALP, iron profile, etc between the *alpha-thalassemia* mutation carriers and non-carriers. A significant elevated level of Total Bilirubin (P = 0.0001), direct bilirubin (P = 0.0004), unconjugated bilirubin (P = 0.0001), SGPT (P = 0.005), SGOT (P = 0.005), ALP (P = 0.008) were observed among the alpha-thalassemia mutation carriers than non-mutant (Table 2).

**Table 2:** Comparison of biochemical parameters in the study population according to mutation profile.

| **Biochemical parameters/Mutations** | **HBA mutant (n = 40)** | **HBA normal (n = 220)** | **P-value** |
| --- | --- | --- | --- |
| Total Bilirubin (mg/dl) | 1.71±0.75 | 0.78±0.46 | **0.0001** |
| Direct Bilirubin (mg/dl) | 0.33±0.19 | 0.2 ±0.14 | **0.0004** |
| Unconjugated Bilirubin (mg/dl) | 1.39 ±0.68 | 0.58 ± 0.39 | **0.0001** |
| SGPT (IU/L) | 54.24±34.34 | 37.01 ± 17.71 | **0.01** |
| SGOT (IU/L) | 47.67±20.24 | 36.93 ± 16.59 | **0.005** |
| ALP (IU/L) | 153.23±103.7 | 101.63 ± 38.12 | **0.008** |
| TOTAL PROTEIN(g/dl) | 7.35±1.02 | 7.34 ± 0.86 | 0.97 |
| ALBUMIN(g/dl) | 4.08±1.05 | 4.10 ± 0.65 | 0.90 |
| TOTAL IRON (µg/dl) | 120.01±57.79 | 118.5± 55.01 | 0.90 |
| UIBC (µg/dl) | 214.16±90.16 | 215.44 ± 94.13 | 0.95 |
| TIBC (µg/dl) | 334.17±90.60 | 329.83 ± 117.36 | 0.83 |
| TSAT (%) | 37.05±17.33 | 36.70 ± 17.54 | 0.92 |
| FERRITIN (ng/ml) | 131.6 | 90.6 | 0.1 |

### Genotype to phenotype correlation

We compared the severity of anemia based on Haemoglobin status between the alpha-thalassemia mutation carriers and non-carriers among our study cohort and we did not find any statistically significant differences (Table 3). Next, on comparing the organomegaly based on HBA gene 3.7 kb or 4.2 kb deletion mutation status, neither the spleen nor liver was correlated with either HBA 3.7 kb or 4.2 kb deletion mutation status. Approximately 85% of *HBA* mutation carrier organomegaly was not present in comparison to 58.6% of the non-carriers (P = 0.0028), suggesting organomegaly may be related to other mutations in *HBA* gene except 3.7 kb and 4.2 kb deletion mutation (Table 4).

**Table 3:** Distribution of HBA gene mutations according to the severity of anemia among the study cohort (n = 260)

| **Study Group** | **Male (n = 123)** |  | **Female (n = 137)** |  | **Total (n = 260)** |  |
| --- | --- | --- | --- | --- | --- | --- |
| **Severity of anemia** | HBA mutant | HBA non-mutant | HBA mutant | HBA non-mutant | HBA mutant | HBA non-mutant |
| **Mild (>10g/dl)** | 3 (15.8%) | 6 (5.8%) | 3 (14.3%) | 18 (15.5%) | 6 (15%) | 24 (10.9%) |
| **P-value** | 0.1392 |  | 0.8854 |  | 0.4583 |  |
| **Moderate (8-10g/dl)** | 16 (84.2%) | 85 (81.7%) | 16 (76.2%) | 77 (66.4%) | 32 (80%) | 162 (73.6%) |
| **P-value** | 0.7956 |  | 0.3789 |  | 0.3968 |  |
| **Severe (6.6-7.9g/dl)** | 0 | 13 (12.5%) | 2 (9.5%) | 21 (18.1%) | 2 (5%) | 34 (15.5%) |
| **P-value** | NA |  | 0.3424 |  | 0.0965 |  |
| **Very severe (<6.5g/dl)** | 0 | 0 | 0 | 0 | 0 | 0 |
| **P-value** | NA |  | NA |  | NA |  |
| **Total** | **19 (15.44%)** | **104 (84.56%)** | **21 (15.33%)** | **116 (84.67%)** | **40 (15.38%)** | **220 (84.62%)** |

Mild anemia: Hb >10g/dl; Moderate anemia: Hb 8-10g/dl; Severe anemia: Hb 6.6 – 7.9g/dl; Very severe anemia: Hb < 6.5g/dl

**Table 4:** Distribution of HBA gene mutations according to Organomegaly among the study cohort (n = 260)

| **Study group** | **Male (n = 123)** |  | **Female (n = 137)** |  | **Total (n = 260)** |  |
| --- | --- | --- | --- | --- | --- | --- |
| **Organomegaly** | HBA mutant | HBA non-mutant | HBA mutant | HBA non-mutant | HBA mutant | HBA non-mutant |
| **Only spleen present** | 3 (15.8%) | 17 (16.3%) | 0 | 18 (15.5%) | 3 (7.5%) | 35 (15.9%) |
| **P-value** | 0.9518 |  | 0.1505 |  | 0.1772 |  |
| **Only liver present** | 0 | 12 (11.6%) | 2 (9.6%) | 12 (10.4%) | 2 (5%) | 24 (11%) |
| **P-value** | 0.2561 |  | 0.9090 |  | 0.2647 |  |
| **Both present** | 1 (5.2%) | 15 (14.4%) | 0 | 17 (14.6%) | 1 (2.5%) | 32 (14.5%) |
| **P-value** | 0.2972 |  | 0.1640 |  | 0.0663 |  |
| **None present** | 15 (79%) | 60 (57.7%) | 19 (90.4%) | 69 (59.5%) | 34 (85%) | 129 (58.6%) |
| **P-value** | 0.0900 |  | **0.0149** |  | **0.0028** |  |
| **Total** | **19 (15.44%)** | **104 (84.56%)** | **21 (15.33%)** | **116 (84.67%)** | **40 (15.38%)** | **220 (84.62%)** |

## DISCUSSION

In this study we have evaluated 260 cases with microcytic anemia with normal Hb-HPLC and decreased iron levels were screened for two common alpha-thalassemia deletion mutations, -3.7kb and -4.2kb in the *HBA* gene. Using the GAP-PCR technique, we were able to detect 40 patients having either 3.7kb or 4.2kb deletion mutations, which represents ~15.38% of the patient population. The 3.7kb deletion mutation was the commonest determinant in this study, which represents 91.8% of the mutations identified. Our study agrees with other Indian studies, where -3.7kb deletion was also the commonest one (Sankar, 2006; Nadkarni, 2007; Sharma, 2015). The 3.7kb or 4.2kb deletion mutations were present among 35.2% of microcytic anemia cases in North India (Sharma, 2015). Borges et al reported that alpha-thalassemia due to a 3.7kb deletion mutation is an important cause of microcytosis and hypochromia in individuals without anemia in the southeastern Brazilian and European population (Borges, 2001; Sivera, 1997).

In our study, we have identified a statistically significant increase in Hb, total bilirubin, direct bilirubin, unconjugated bilirubin, SGPT, SGOT, and ALP levels among the deletional mutation carriers than non-carriers. Our data is consistent with a report, where the author reported that the Hb level is significantly higher among the carriers than non-deletional mutation carriers (Vijian, 2023). In contrast to our findings, there is an increase in MCH value among the mutants (Vijian, 2023). Our study demonstrated that the level of total, direct, and unconjugated bilirubin was high among the deletion mutation carriers which is consistent with another report from north India (Panigrahi, 2006). The authors reported that the severity of jaundice was higher in thalassemia intermedia cases with HBA gene mutation (Panigrahi, 2006).

In conclusion, the two common deletion mutations in the *HBA* gene may explain ~15.38% of microcytic anemia cases in West Bengal and several biochemical factors were correlated with the mutation status.

## Data Availability

The data that support the findings of this study are available from the corresponding author upon request.

## Acknowledgement

The authors thank the patients who participated in the study.

## Data Availability Statement

### Role of the Authors

SB, SG, RD, and AB, were responsible for the concept, study design, sample collection, experimental work, and manuscript preparation. SNB was responsible for sample collection, clinical diagnosis, and manuscript preparation. AS, and SDG were responsible for sample collection. All authors read the draft, provided their inputs, and agreed on the final version of the manuscript.

### Ethics Approval

All procedures performed in studies involving human participants were in accordance with the ethical standards of the Nil Ratan Sircar Medical College & Hospital, Kolkata, India. The Ethics Committees (Nil Ratan Sircar Medical College & Hospital, Kolkata, India) of the abovementioned Institutes approved the study protocol. Informed consent was taken as per guidelines of the Indian Council of Medical Research, National Ethical Guidelines for Biomedical and Health Research involving human participants, India.

## Conflict of Interest

The authors declare that they have no conflict of interest.

### Informed Consent

Informed consent from all the participants was received before clinical data and sample collection.

## References

Borges E, Wenning MR, Kimura EM, Gervásio SA, Costa FF, Sonati MF. High prevalence of alpha-thalassemia among individuals with microcytosis and hypochromia without anemia. Braz J Med Biol Res. 2001 Jun;34(6):759–62. doi: 10.1590/s0100-879x2001000600009. PMID: 11378664.

Chong SS, Boehm CD, Higgs DR, Cutting GR. Single-tube multiplex-PCR screen for common deletional determinants of alpha-thalassemia. Blood. 2000;95(1):360–362.

Dash S. Beta thalssemia trait in the Punjab (North India). Br J Haematol. 1985; 61:185–186.

Husna N, Handayani NSN. Molecular and Haematological Characteristics of alpha-Thalassemia Deletions in Yogyakarta Special Region, Indonesia. Rep Biochem Mol Biol. 2021 Oct;10(3):346–353. doi: 10.52547/rbmb.10.3.346. PMID: 34981010; PMCID: PMC8718782.

Inusha Panigrahi, Manoranjan Mahapatra, Rajat Kumar, Guresh Kumar, Prakash Choudhry Ved & Renu Saxena (2006) Jaundice and alpha gene triplication in beta-thalassemia: Association or causation?, Hematology, 11:2, 109–112, DOI: 10.1080/1024533050069882

Karakas Z., Koc B., Temurhan S., Elgun T., Karaman S., Asker G., Gencay G., Timur C., Yildirmak Z.Y., Celkan T. Evaluation of alpha-thalassemia mutations in cases with hypochromic microcytic anemia: The Istanbul perspective. Turk. J. Hematol. 2015; 32:344. doi: 10.4274/tjh.2014.0204.

Misra RC, Ram B, Mohapatra BC, et al. High prevalence and heterogenicity of thalassemias in Orissa. Indian J Med Res. 1991; 94: 391–394.

Nadkarni A, Phanasgaonkar S, Colah R, Mohanty D, Ghosh K. Prevalence and molecular characterization of alpha-thalassemia syndromes among Indians. Genet Test. 2008;12(2):177–180. doi:10.1089/gte.2007.0080

Sankar VH, Arya V, Tewari D, Gupta UR, Pradhan M, Agarwal S. Genotyping of alpha-thalassemia in microcytic hypochromic anemia patients from North India. J Appl Genet. 2006;47(4):391–5. doi: 10.1007/BF03194650. PMID: 17132905.

Sen R, Chakrabarti S, Sengupta B, et al. Alpha-thalassemia among tribal populations of Eastern India. Hemoglobin. 2005;29(4):277–280. doi:10.1080/03630260500310711

Sharma RS, Parekh JG, Shah KM. Hemoglobinopathies in western India. J Assoc Physicians India. 1963; 11: 969–973.

Sharma M, Pandey S, Ranjan R, Seth T, Saxena R. Prevalence of alpha thalassemia in microcytic anemia: a tertiary care experience from north India. Mediterr J Hematol Infect Dis. 2015 Jan 1;7(1):e2015004. doi: 10.4084/MJHID.2015.004. PMID: 25574363; PMCID: PMC4283920.

Sivera P, Roetto A, Mazza U, Camaschella C. Feasibility of molecular diagnosis of alpha-thalassemia in the evaluation of microcytosis. Haematologica. 1997 Sep-Oct;82(5):592-3. PMID: 9407728

Vijian D, Wan Ab Rahman WS, Ponnuraj KT, Zulkafli Z, Bahar R, Yasin N, Hassan S, Esa E. Gene Mutation Spectrum among Alpha-Thalassaemia Patients in Northeast Peninsular Malaysia. Diagnostics. 2023; 13(5):894. 10.3390/diagnostics13050894

